# Experience(s) of the medical profession: A qualitative study using narrative facilitators

**DOI:** 10.1101/2021.09.30.21264334

**Authors:** Céline Bourquin, Sandy Orsini, Friedrich Stiefel

**Affiliations:** Psychiatric liaison service, Lausanne University Hospital and University of Lausanne, Lausanne, Switzerland

## Abstract

Physicians’ narratives are means to approach and comprehend their experiences. They reveal the practice of medicine, and inform about the physicians’ embedment in their work and the healthcare context. This study aimed to examine physicians’ experience by means of interviews based on “narrative facilitators”, which goal is to encourage storytelling and to support the narrative process. Core stories were elaborated on the key aspects that emerged from the analysis. Thirty-three physicians participated in the study. The findings showed a focus on the transformations of a profession, the need for physicians to adapt in terms of role and status, and their withstanding of conflicting projections from the public and patients. The institutional context was described as darkness in which physicians are moving. When reacting to the quotes from their peers, participants showed a variety of un-patterned stances with regard to different aspects of medicine and the medical profession. Findings also indicated that as narrators, physicians may have particular difficulties accessing their inner world. Disenchanted physicians are not beneficial, neither for the patient nor for the health care system, and their feeling of being worn out may do harm and is certainly negatively affecting themselves and their families.

## Introduction

Over 10 years ago, in a Lancet article entitled ‘The suffering of physicians’, Cole and Carlin reminded that the patient is not the only “whole person” in the consulting room, and that physicians also suffer from the “dehumanization” of modern medicine (1). The issues they addressed, notably in relation to the clinical encounter, have meanwhile become of utmost importance (2,3). To allow vulnerable physicians to recover meaning and to avoid burnout, Cole and Carlin recommended “to respect” physicians’ stories. This evidently requires both that physicians tell their stories and that somebody listens (1).

Whether physicians suffer or not, their narratives are means to approach and comprehend their lived experiences (4,5). Such narratives reveal the practice of medicine, but also inform about the physicians’ embedment in their work and the healthcare context. They are a precious resource offering insights into how physicians perceive their professional and personal challenges (5,6). Moniz et al. pointed out in a recent article on systems flaws identified by physicians in written reflective narratives that “narratives is a valuable mode of self-expression in medicine and that, by engaging with narrative, we can come to grasp the meanings and interpretations that individual storytellers ascribe to their experiences” (7).

The overwhelming majority of studies exploring physicians’ experiences focuses on how they cope and struggle with issues such as budget constraints and workload or anxiety, depression, and burnout (8,9). Alongside these studies, the issues of identity as physician (physicianhood) (10,11), professional socialization (12) as well as professional identity formation during medical training have received attention (13,14). More rarely, the psychological functioning of physicians has been examined, for instance by investigating their defense mechanisms (15). Actually, we do not know much about the way the health care institution, patients’ and public’s expectations towards medicine and physicians, or society’s dominant discourses impact physicians’ experiences.

This study was triggered by an observation made during a qualitative investigation aiming to approach the perspective of physicians on communication with and care of dying patients. This observation of “what was silenced” in focus group discussions can be summarized as follows: physicians did neither spontaneously talk about their experiences nor about contextual factors of their daily work. We therefore decided to proactively invite them to tell their stories about their professional experiences and to examine how they are affected by factors related to their inner (psychological) and outer (contextual) world.

## Methods

The study was designed as an exploratory qualitative study to examine physicians’ lived experiences by means of interviews based on “narrative facilitators”.

### Data collection

We have developed facilitators of narratives as a method to empirically address the factors, which might impact physicians’ experiences. The facilitators of narratives rely on techniques inspired by visual sociology (use of photographs as narrative support [photo-based story]) (16), clinical psychology (Thematic Apperception Test [blurred video sequences]) (17) and on purpose-designed techniques (use of press articles and quotes from peers [press-book and quotes from biographies/narrative accounts of the experience of being a physician]). This method aims to encourage storytelling and to support the narrative process on experience. Put differently, narratives were produced as an artefact of research, but with prompts (our facilitators) aiming at facilitating and supporting the narratives without being overly constraining.

As shown in Table 1, the interview guide consisted of four distinct types of facilitators of narratives. Lasting about 90 min, the interviews were conducted in 2016-2017 by the first and the second author and audiotaped. They were analyzed directly afterwards, but the drafting of the manuscript was delayed due to an overlap with other projects.

**Table 1.** Interview guide.

| Investigated aspects | Narrative facilitators | Instructions |
| --- | --- | --- |
| Dominant discourses on medicine and the medical profession | **Press-book:**<br>8 booklets with articles about nowadays hospitals, physicians' roles, medical malpractice, heroic physicians, evolution and revolution in medicine | What kind of a story do all these booklets tell about medicine and physicians? How do you feel about the story (what is your reaction)? |
| Contextual factors related to the medical institution and apparatus | **Photo-based story:**<br>8 *ad hoc* designed photographs featuring a working day of two physicians in Western university hospitals (from their arrival to the moment they leave the hospital at the end of their shift, seeing patients, walking in the hospital, attending meetings, etc.) | What do you know about the man featured in the photos and what is he doing? How do you know that? |
| Contextual factors related to physicians' experience of their work and relationship with peers | **Quotes from physicians' biographies/narrative accounts of the experience of being a physician:**<br>10 plasticized sheets addressing topics like medicine as a vocation, physician burnout, empathy, etc. | To what extent (10-point analogue scale) do you agree with this quote? Why do you agree/disagree? |
| Physicians' inner world | **Short (2 min) video sequences:**<br>4 blurred video sequences without sound from two documentary films on the profession of medicine, which feature a physician doing administrative duties, a physician talking to a bedridden patient, a group of physicians, and a physician alone in front of an elevator turning in circles | What do you see in this sequence? What determined what you say? |

### Participants

In continuation of our study on the perspective of physicians on end-of-life communication and care, but without a specific hypothesis or question with regard to sampling, we conducted interviews with chief residents and senior staff members working in geriatrics, palliative care, oncology and internal medicine at Lausanne University Hospital.

Participating physicians provided written informed consent. The Human Research Ethics Committee of Vaud exempted the study from approval.

### Data analysis

The analytic approach was specific for each facilitator. It may seem to be a non-obvious choice, but it was a way to stay close to the raw narrative data and to the four different aspects that we examined (dominant discourses, the institutional context in which physicians are working, their socialization with peers, and their psychological state). The data also reflected dimensions that were external or internal to the physicians. In other words, we used different techniques of analysis to differentiate between introspective data (related to the inner world of the physicians) and more extrospective data.

Specifically, the dominant stories outlined by physicians based on the press-book, were analyzed with thematic analysis (18). Patterns of stories were identified and characterized through an iterative, open-coding process.

Narratives of physicians related to the photo-based story of a hospital physician were examined with a photo-elicitation analysis (19). The analysis focused on (i) what we aimed to show with each photograph (photo composition) and what physicians told about it, (ii) and what else the physicians saw in the pictures.

With respect to the quotes, which were evaluated on a 10-point analogue scale, we used a statistical-based approach. Latent class analysis on polytomous variables served to detect any potential latent classes among our sample, which may have a similar stance with respect to the quotes (20). To do so we employ the poLCA package in R, which maximizes the log-likelihood of each latent class model using EM and Newton-Raphson algorithms (21). The focus of the analysis for the first three facilitators was on the told and telling. In contrast, the narrative material produced in relation to the video sequences was analyzed by means of the cues provided by the individual storytellers (physicians), which provided insight into their concerns and interests, and allowed us to explore their self-representations.

Physicians’ narratives related to the facilitators were distinct but deeply interrelated. In the results, we report on the core stories, elaborated on the key aspects that emerged from the analyses.

### Reflexivity

Two investigators analyzed the data (CB and SO). They met regularly with the third investigator (FS) to review the analyses. The different perspectives and backgrounds of the investigators enabled sound discussions of findings and interpretations. CB is a social science researcher (anthropologist) embedded in a medical service for years, SO is a psychologist, and FS a senior consultation-liaison psychiatrist.

## Results

33 physicians of Lausanne University Hospital participated in the study (13 were female). Five physicians worked in geriatrics, six in palliative care, 10 in oncology, and 12 in internal medicine.

As part of the reported core stories, we provide room for physicians’ voices by means of interview excerpts. Individual physicians are identified by an alphabetical letter and their disciplinary affiliations are indicated.

### The changing profession of medicine

The main story told by physicians, elicited by *press discourses*, was that of a profession in transformation, and of a changing social status. To practice medicine nowadays requires new qualities and competences: clinical expertise is not enough; it has to be completed by management skills, particularly to contain cost. New dimensions have to be taken into account to respect a holistic approach of the patient, as promoted, for instance, by the bio-psycho-socio-spiritual model of care. Physicians are no longer the Gods in white; they make errors for which they are taken accountable. This has an impact on the clinical relationship and may undermine trust.

In some versions of this story, the transformation of the profession of medicine, especially related to scientific progresses and the introduction of new technologies, is associated nonetheless with a dehumanization of care: performance and skills dominate over relational and humane competence. More or less directly linked to these changes, some physicians perceived an ambivalent stance of the public towards medicine, oscillating between fear and fascination, and blind trust and contempt.

> […] we are in a time when we [medicine in general] are losing a little bit of our identity, and we are also losing ground […] we attempt by different means either to regain prestige or to demonstrate in every possible way that we are critical of ourselves or that we are incredible with regard to technological advances […] This is a moment of identity crisis, whereas medicine was always rather esteemed, and occupied an important place. I consider that as a shift […] (N, palliative care)
>
> […] the physician of the beginning of the last century, as we imagine him, was never challenged, people trusted him. I do not say it was the good old time (laughs) […] But the [current] system, according to me, generates a lot of fear among the public, and also hope, fascination (H, palliative care)

Certain physicians colored this narrative of a changing medicine positively. They saw a rather triumphant medicine, capable of inspiring trust due to advances in science and technology, quality training objectives, and skills demonstrated by physicians. This medicine offers endless possibilities for the medically ill, who responds to this offer and engages with medicine.

> […] I think people are increasingly interested in their health, and sometimes in an obsessional manner, but generally I consider this is a rather positive evolution. People ask themselves questions like “how can I live healthier?” […] “how should I choose a hospital”? “Where will I get the best care?” […] I consider this rather positive (X, oncology)

### Meanwhile we are used to this

Passing the doors of the hospital, the stories, elicited by the *photos*, revealed a more pragmatic thinking. Physicians were described as very busy, overworked, tired, and even exhausted, lacking energy, due for instance to administrative work, and feeling isolated.

> […] I imagine that he is not doing what he wants to do, well he is not in patient care, he is not related to another person, he is just doing paper work […] and that’s not the most interesting part of the job (H, internal medicine)
>
> […] did we choose medicine for that [being in front of a computer]? No, but if you don’t do it, you don’t get paid. We are fewer and fewer [physician shortage], it’s a vicious circle (L, internal medicine)
>
> […] a day at the hospital […] we arrive, we see patients […] and we can observe how late we are already (laughs), and we try to do paper work. Once everybody is back home, we still make phone calls, look at the x-rays, prepare the next day, then we also have meetings lasting for hours with liaison nurses, who have requests and who make you fill in 15 different forms. And then, finally, night comes, you are tired, you take off the white coat and you go home. That’s how it feels (X, oncology)

The global (the hospital) and local (the office) working space was viewed as not welcoming, impersonal and even depersonalizing. Daylight does not penetrate. An environment on which physicians have not much to say or to decide upon.

> […] You can see that on strategic places, they put on some stuffs to give a modern look, with an attempt to bring in some life and decoration in the hospital […] (L, internal medicine)
>
> […] the offices of the nurses and of the technicians are refreshed first, and only then those of the physicians. With regard to ergonomics, etc., physicians always come last […] I even saw physicians working just beside the hood for chemotherapy […] there has been this protest for those who work in a place without windows, they can take more holidays. Well, the problem is you have to participate in these meetings [to organize claims] you need time, but we don’t have time, and thus people [administrators] consider that it’s not important to us. We don’t have that lobby […] we have a lot of things which disturb us, and we go along with them (L, internal medicine)

### Being a physician is an individual experience

When one addresses the shared (or unshared) perspectives of physicians on different issues specifically related to the practice and vision of the profession of medicine, the previous relative homogeneity in participants’ narratives disappear. Indeed, we could not find evidence for the presence of latent classes. In other words, the physicians did not cluster into groups, which provide same answers and perspectives on some of the questions related to the *quotes*. Some quotes evoked *polarized reactions* illustrating split opinions with regard to specific experiences of medicine. Physicians were divided on issues, such as sharing daily work experiences with loved ones, personal strain related to work or the detachment from emotions. The two following excerpts show the split views on this latter issue:

> […] I got used to these things, so now I have even sometimes the impression to lack compassion, sometimes I do not feel sad enough with regard to things I heard […] I have the impression that this compassion, it has left me, since I have already heard these stories before (Q, oncology)
>
> […] I think I go a little bit further than empathy, I try not to reach compassion, I try not to reach sympathy, but probably I go a little bit further than pure empathy, because this pleases me, it allows me to foster relationships […] (C, internal medicine)

Medicine considered as a calling gave rise to the most *various perspectives*.

> […] the majority [of physicians] they have a sense of calling, otherwise it is not possible to do this job (T, oncology)
>
> […] I think one have to be honest, it [the job of a physician] becomes a job like any other […] if physicians had a sense of calling, there would be no millionaires among them, and they wouldn’t leave to the private sector (E, oncology)

Responses and narratives from physicians were more *homogeneous* when quotes from peers addressed issues, such as medicine as a team effort – a profession with no room for self-gratification –, or medicine as administrative-governed and profit-driven.

> […] sometimes one gets the impression that we have become medical secretaries. The secretaries drop things back to us, and the desk clerk yells at us, since we did not fill in this and this sheet correctly, and radiology calls that they did not get the correct form. You are in front of a pile of documents to fill in for insurance, and for sure then you find yourself as being very dehumanized. Indeed, nobody considers that we are physicians, that we are here to care for patients and not to stack files […] (X, oncology)
>
> […] I hate the administrators, I feel not afraid to say it because it’s true, these people add obstacles to the system, they force us to complete tons of forms and supporting documents […] (M, internal medicine)

### Accessing one’s inner world is difficult

While the *video sequences* aimed to orient physicians to respond to a scene by filling it with their own interest and concerns, they often limited their narratives to a description of the materiality of the elements putted into play, thus doubling in words what was put on the screen. The production of free associations, emotions or fantasies were rather lacking. A small minority of physicians referred though to their own experiences, such as feelings of anonymity or on the contrary stressed the importance of exchange between peers.

> […] Well, its half past four in the hospital [the participant refers to clock, which appears in the sequence], it seems to be in pediatrics or something like this, well, it seems to me I recognize a colleague. This video, mainly, it shows plenty of physicians who are in front of a meeting room, I don’t know if it’s just before or after the meeting and they discuss together and then there is another group of physicians, a smaller one, but still seven or eight, and they discuss together and then they leave, probably to join their offices (O, geriatrics).
>
> […] There is a physician, I think he’s a physician, since he seems to have a pager, let’s say he looks like a physician. He strolls in the corridors, he seems preoccupied […] Well, he seems to think about something […] hands in the pocket, the upper or lower pockets, this way, as someone who thinks, a little bit of psychomotor agitation, which could indicate that he either waits for the elevator or he is impatient […] and then, he seems lost in his thoughts, since the nurse walks by, and they don’t interact (I, geriatrics)

## Discussion

We conducted this study in a nowadays medical context with negative physical and mental health consequences and a gradual decline of satisfaction among physicians (2,8,22–26). Besides these studies assessing specific aspects of health and job satisfaction of physicians, naturally occurring narratives by physicians are mostly found in dedicated books or in narrative sections of medical journals (5,7,27). In the meantime, the SARS-CoV-2 pandemic has shown that in an extraordinary context and in situations of crisis, physicians are strongly encouraged to tell their experiences about the practice of medicine (28). This study is part of an effort to provide room for their voices and stories even, or especially, in “ordinary times” (29–31).

The scope of this exploratory study was broad. Given the observation that physicians’ experiences with regard to their inner (psychological) and outer (contextual) world were not easy to elicit, we attempted to encourage their narratives by means of facilitators linked to different dimensions. Among them were – following a centripetal process – dominant discourses about medicine and physicians, the institutional context in which physicians are working, the peers with whom they socialize, and finally their psychological state. The interest physicians showed for the study during recruitment, their motivation when engaging with the facilitators of narratives and the pleasure they took to talk about their “view of the world”, somehow confirm us in our approach. Physicians want to tell their stories, and the facilitators contributed to support the narrative process. Compared to the more elaborated and structured published stories, physicians seemed inclined to share experiences without filtering them through a process of rationalization or courtesy.

The findings show that the participants’ narratives focus on the transformations of a profession, the need for physicians to adapt in terms of role and status, and their withstanding of conflicting projections from the public and patients. What hurts is the negative impact of these transformations on the clinical work, which remains as their core business more and more difficult to protect, and the relationship with the patient, even more so since physicians consider facing a public who stares at the hospital with mixed and ambivalent feelings. Moreover, the impression prevails that they are left on the side of the road, deprived of the agency to shape their destiny. The positivistic narratives of a minority of participants, applauding scientific progress and achievements of modern medicine, cannot evacuate the rather low morale physicians are demonstrating when looking at their professional lives; a result, which is in line with studies conducted in other health care settings (32,33). In a recent article, Monitz et al. (7) identified sources of tension and distress for physicians and showed the ways physicians are challenged by system flaws, and try to maintain resilience and to “live up to the ideals of the profession.”

Echoing the latter observations, the institutional context was metaphorically described as darkness in which physicians are moving, again with a certain feeling of bearing their fate. Tired and worn out, and somehow resigned, however, they seem gotten used to play the game.

On the other hand, physicians remained quite singular and demonstrated an independent mind, when reacting to the quotes of their peers, showing a variety of un-patterned stances with regard to different aspects of medicine and the medical profession. This finding underlines the need for an approach, if interventions are considered, which is tailored to the particular situation and needs of physician; this is also well known from mentoring and other efforts to support physicians during their development (34). Indeed, physicians are shaped not only by their socialization, but also by biographical events and their private context, which influence career choice, but also career constancy (35).

While physicians’ opinions related to “medical matters” appear to be individually formed, leaving room for singularity in the way the profession is conceived, and easily expressed, it seems more complicated for them to access the underlying psychological dimensions associated with their lived experiences. Indeed, the exercise to elaborate on the video sequences putting into play the physician’s office work, the clinical encounter, the relationship with peers and himself/herself, was considered as a difficult task. As a consequence, physicians tended to double what was projected in the scenes, and the material provoked limited resonance with their inner world. This last result may not be so surprising, since the promotion of reflection and psychological insight are rather rare during the under- and postgraduate medical education, and a lot of emphasis is put on gaining knowledge and learning how to applicate knowledge.

How can we understand these narratives? Two main observations seem to be at the core of the lived experiences of the physicians.

First, physicians deplore an atomization of their professional identity: multi-tasking leads to switching identities, multiple injunctions drives them away from their core business and the tasks of fulfilling conflicting goals operate as double bind messages and provoke a feeling of alienation. This professional atomization, together with their singularity when it comes to conceive medical matters, seems to hamper an effective unionization of the profession, which could defend physicians’ interests. Their interests are perceived as disregarded, according to the narratives, and lead to a feeling of a loss of agency. Atomization, and the difficulty to stick together and defend the issues at stake, feels like a continuation of what physicians experienced during the socialization process in medical school, which is after all characterized by high competiveness and selection, shaping their identities way after the final exams (29). This first result is in sharp contrast with the efforts made to welcome patients in the hospital, to respond comprehensively to their needs, to perceive them in their individuality and to promote patient-centered care, shared decision-making, empowerment, partnership, and expression of complaints, which are regarded as a learning experience for the health care personnel (36). The question thus arises: should not we care more for physicians?

Second, physicians seem to endure their work experiences without situating themselves. This feeling of being “lost in medicine” is illustrated by the difficulties to elaborate on the video material, which invites to freely associate and to establish links with own experiences, nurtured not only by cognitions, but also emotions. Here again, the recent efforts made to take into account the subjectivities of patients by training physicians in medical psychology and communication (37,38) and to associate patients to better understand and integrate their perspective in the care (39), stand in sharp contrast to what has been undertaken to help physicians to access their inner world and to situate them reflectively in their work context. The introduction of Balint groups, individual supervision, and other interventions such as the so-called Osler groups or reflective training for clinicians are rather the exception than the rule (34,40,41). Moreover, many of these offers are accessible on a voluntary basis, and the impression prevails that they are used by the motivated few who need it least. The question thus arises: what can be done for physicians?

We consider that pour study has the potential to contain elements, which contribute to answer the two previous questions. First, the experience of a loss of agency, of an atomization of the physician’s identity and of isolation clearly underlines that physicians need help. It is not good enough to recognize and treat physician burnout and to regularly assess job satisfaction. One of the reasons why the lived experience of physicians is not really taken into account might lie in the lack of conceptual framework to tackle the question of how to help them, and this brings us to the second question we raise.

What for patients has become self-evident – agency, respect for their singularity and the expectation to be in good hands – is less natural for physicians; the reasons may be manifold (e.g., prosocial identity of physicians, the context of sever suffering, a sense of duty or an attitude of lone ranger). However, what matters is that a conceptual framework of how to “care for physicians” is lacking. Such as framework cannot be based on the psychological or cognitive aspects of being a physician today, it has to include their affective and lived experience. Psychological approaches to access the inner world of physicians are precious as are emerging reflexivity training but they need to be complemented by methods which access the experiences revealed in this study, which physicians have a tendency to hide or neglect. In conclusion, even if disenchantments of physicians might not be a completely new observation (33), their underlying reasons should be addressed. Disenchanted physicians are not beneficial, neither for the patient nor for the health care system, and their feeling of being worn out may do harm and is certainly negatively affecting themselves and their families. Physicians, as patients, need respect and support, and deserve to be given the same attention – as stated by Gorlin and Zucker already in 1983 (42).

## Data Availability

The data that support the findings of this study will be available in Zenodo, when the article will be published

## Declarations

### Ethics approval

The Human Research Ethics Committee of Vaud exempted the study from approval.

